# SARS-CoV-2 shifting transmission dynamics and hidden reservoirs limited the efficacy of public health interventions in Italy

**DOI:** 10.1101/2020.12.16.20248355

**Authors:** Marta Giovanetti, Eleonora Cella, Francesca Benedetti, Brittany Rife Magalis, Vagner Fonseca, Silvia Fabris, Giovanni Campisi, Alessandra Ciccozzi, Silvia Angeletti, Alessandra Borsetti, Vittoradolfo Tambone, Caterina Sagnelli, Stefano Pascarella, Alberto Riva, Giancarlo Ceccarelli, Alessandro Marcello, Taj Azarian, Eduan Wilkinson, Tulio de Oliveira, Luiz Carlos Junior Alcantara, Roberto Cauda, Arnaldo Caruso, Natalie E Dean, Cameron Browne, Jose Lourenco, Marco Salemi, Davide Zella, Massimo Ciccozzi

**Author notes:** These authors contributed equally to this article (Marta Giovanetti, Eleonora Cella, Francesca Benedetti, Brittany Rife Magalis).

## Abstract

We investigated SARS-CoV-2 transmission dynamics in Italy, one of the countries hit hardest by the pandemic, using phylodynamic analysis of viral genetic and epidemiological data. We observed the co-circulation of at least 13 different SARS-CoV-2 lineages over time, which were linked to multiple importations and characterized by large transmission clusters concomitant with a high number of infections. Subsequent implementation of a three-phase nationwide lockdown strategy greatly reduced infection numbers and hospitalizations. Yet we present evidence of sustained viral spread among sporadic clusters acting as “hidden reservoirs” during summer 2020. Mathematical modelling shows that increased mobility among residents eventually catalyzed the coalescence of such clusters, thus driving up the number of infections and initiating a new epidemic wave. Our results suggest that the efficacy of public health interventions is, ultimately, limited by the size and structure of epidemic reservoirs, which may warrant prioritization during vaccine deployment.

## Main text

On December 31^st^ 2019, the World Health Organization (WHO) China Country Office was informed of pneumonia cases of unknown aetiology detected in Wuhan City, Hubei Province^1,2^. By January 11^th^ – 12^th^ 2020, Chinese authorities identified a novel single stranded, positive-sense enveloped RNA *Betacoronavirus*, with genome of 30,000 nucleotides in length, belonging to the *Coronaviridae* family, related to the severe acute respiratory syndrome coronavirus (SARS-CoV) that caused a global outbreak in 2002–2004 ^3^. Initially named nCoV-2019 (novel Coronavirus 2019), the virus likely emerged from several recombination events in bats and pangolins ^4^, and was subsequently introduced in the human population through zoonotic transmissions ^1,5^; it was later renamed SARS-CoV-2, and recognized as the etiologic agent of Coronavirus Disease 2019 (COVID-19) ^6^. Epidemiological investigations and phylogenetic analysis promptly confirmed airborne SARS-CoV-2 human-to-human transmission ^3,7^. Following its worldwide spread, the WHO declared the outbreak as a Public Health Emergency of International Concern on January 30^th^, 2020, and a pandemic on March 11^th^, 2020. As of December 16^th^, 2020, SARS-CoV-2 has spread to 216 countries with nearly 74 million confirmed cases and over 1.6 million fatalities ^8^.

Italy was one of the first and most affected countries in the world. By October 31^st^ 2020, the Italian Ministry of Health and the Civil Protection Department reported 1.38 million total SARS-CoV-2-related cases, and 49,261 deaths ^9^. The first confirmed imported cases dated back to January 30^th^ 2020 when two tourists from Wuhan, China, were tested positive for SARS-CoV-2 in Rome (**Figure 1A**). On February 17^th^ 2020, the Italian government confirmed the first locally acquired case in a small city in Northern Italy (Codogno, Lombardy region) ^10^. Three days later, the first COVID-19-related death in Italy, a 78-year old male, was reported in the city of Padova. As the epidemic quickly spread throughout the country, establishing Italy as one of the major SARS-CoV-2 hotspots ^11^, the Italian government declared a Public Health Emergency of National Importance, enabling the introduction of restriction measures to limit new infections ^12^. In the effort to flatten the epidemic curve, Phase I lockdown measures were first introduced on March 7^th^ – 8^th^ 2020 in 11 municipalities of Northern Italy, where most cases had occurred, and extended by March 11^th^ to the whole country (**Figure 1A**). Described as the largest lockdown in the history of Europe ^13^, citizen mobility was restricted, except for “well grounded” work- or health-related reasons. A universal mask mandate was required at all times outdoors. Schools, university activities, public/cultural events, and sport competitions were also suspended nationwide, as well as non-essential commercial activities. Borders with other states were closed, and within the country public transport was limited or shut down.

**Figure 1.**
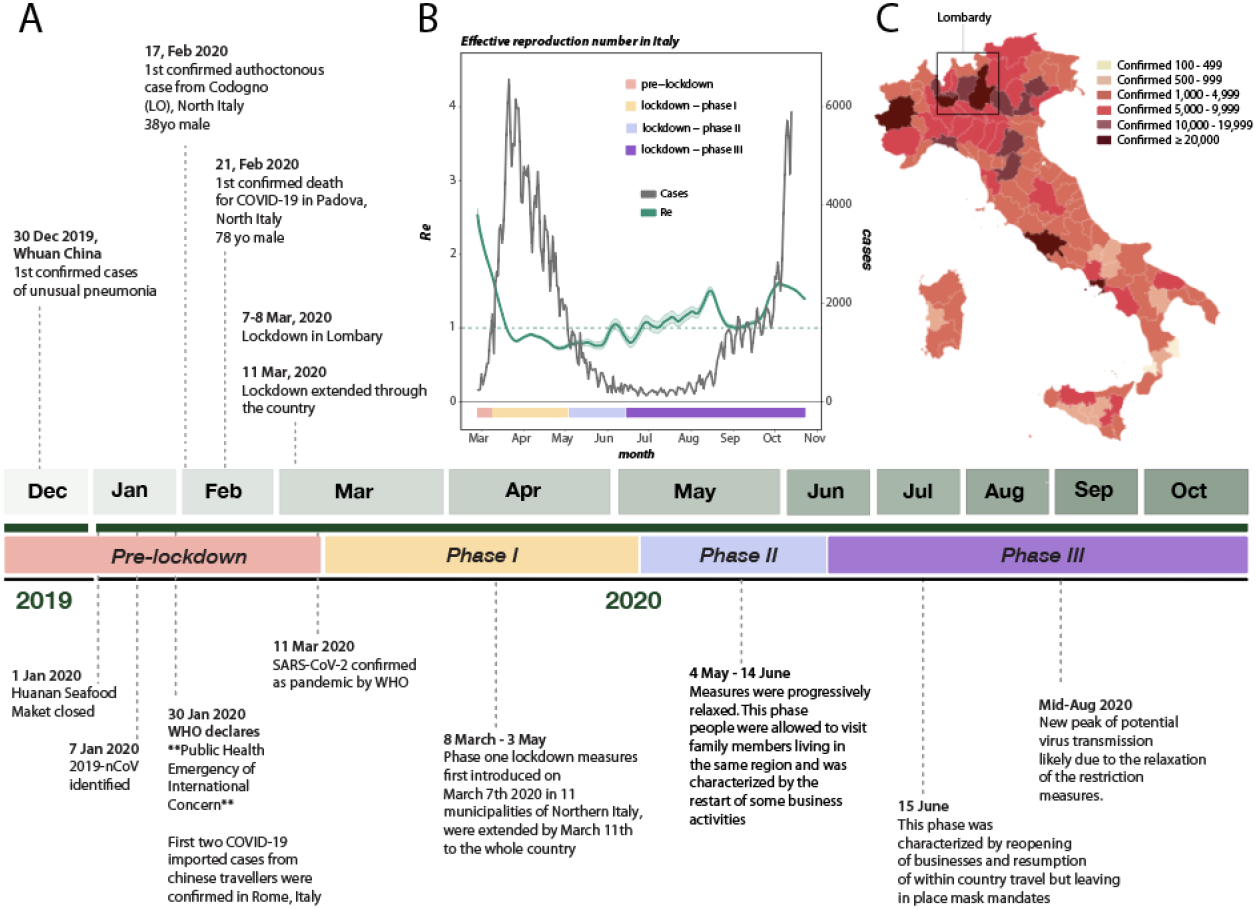
History of SARS-CoV-2 epidemic in Italy. **A**) Timeline of key events following the first confirmed cases of SARS-CoV-2 infection in Italy. **B**) Epidemic curve showing the progression of reported daily viral infection numbers in Italy from the beginning of the epidemic in March (black) and changes in *Re* estimations in the same period (green), with lockdown phases indicated along the bottom. **C**) Map of cumulative SARS-CoV-2 cases per 100,000 inhabitants in Italy up to Oct 2020.

As daily viral infection numbers decreased, public health measures were progressively relaxed through a Phase II (May 4^th^), which allowed visits to family members living in the same region and the restart of some business activities, and a Phase III (June 15^th^), which allowed reopening of businesses and resumption of within country travel, but left in place mask mandates and bans on large-scale meetings. A significant slowdown in the number of infections since the beginning of May 2020 (**Figure 1B**) validated the effectiveness of Phase I restrictions.

After a period of seemingly stable epidemic recession, with very few new cases detected between June-August, a new epidemic wave hit the country, resulting in higher incidence than before. Superimposition of the reported epidemic curve and dynamic estimates of the effective reproduction number, *Re*, throughout the three major periods (first wave, recess, second wave) of the Italian epidemic, revealed an interesting pattern (**Figure 1 B**). *Re* provides a measure of the average number of secondary infections caused by a single infected person: a growing epidemic is typically characterized by *Re* > 1, while *Re* < 1 indicates no growth. As expected, *Re* values were estimated to be > 2 at the beginning of SARS-CoV-2 exponential spread in Italy, and quickly fell to values < 1 after the start of Phase one lockdown measures. Yet, between end of June and end of August, through Phase II and III lockdowns, *Re* values showed an oscillating behaviour, with progressively higher peaks (>1), despite the consistently low number of newly detected infections. As infections and hospitalizations began to climb in September, *Re* temporarily decreased close to 1, to increase again by mid-October, just before the beginning of the new exponential growth of infected cases, currently ongoing. Indeed, by October 31^st^, all Italian regions, albeit with different rates were hit by the epidemic (**Figure 1C**). The rapid increase of COVID-19 patients requiring hospitalization during the early months of 2020, as well as *Re* oscillations during the period of epidemic recession, suggest the virus was circulating cryptically among undetected transmission clusters. During this time, there were possibly thousands of mild or asymptomatic infections among undetected (hidden) reservoirs that preceeded each exponential growth phase of each epidemic wave ^14,15^. Indeed, dramatic resurgences in cases after easing stringent public health interventions (i.e., stay-at-home orders) that temporarily curtailed epidemic spread was also observed in several other European countries (e.g., UK, France, and Germany, among others).

To investigate further, we coupled epidemiological data with phylodynamic analysis of 714 viral sequences currently available from Italian patients, sampled between January 30^th^ to October 1^st^, 2020 (see Methods). Viral population dynamics were assessed using non-parametric coalescent estimates of the effective population size (*Ne*) over time (a measure of viral diversity representing the number of diverse genomes contributing to the next generation), given a collection of plausible maximum likelihood (ML) evolutionary histories inferred from viral sequence data. Although distinct patterns could be observed in *Ne* estimates, all reconstructions agree on a rise in *Ne* until the end of March 2020, matching the rise in number of reported cases (**Supplementary Figure 1**). The best-fit model (i.e. the collection of trees with the highest likelihood, *log*L > −49,120) also depicts a steady, continuous decline in *Ne* until October (**Supplementary Figure 1**, pattern A), possibly reflecting the impact of lockdown measures on the viral population. As *Ne* is related to viral genetic diversity, this pattern may indicate that, despite the rapid rise of cases in late summer, the viral population maintained lower diversity relative to the earlier months of the epidemic. This is consistent with a reduction of viral importations, likely resulting from global public health intereventions such as travel bans. Two alternative patterns inferred from trees similar in likelihood value to the best-fit model, show either a similar downward trend followed by a pronounced increase in *Ne* between September and October (pattern B), or a slower but steady increase in *Ne* between April and October (pattern C). Both reconstructions are in agreement with an increase of viral *Ne*, corresponding with an exponential increase of SARS-CoV-2 infections during the second epidemic wave. Together, with the inferred oscillations of *Re* values following the first epidemic wave, the analyses suggest the persistence of complex transmission dynamics throughout the epidemic recession period, involving undetected asymptomatic or mildly affected individuals. Even considering the *Ne* values inferred from ML trees with lower likelihood, we arrive at an analogous conclusion – overall reduction in *Ne* after April but repeated fluctuations throughout recession and second epidemic wave (**Supplementary Figure 1**, black curves).

Longitudinal comparison of SARS-CoV-2 dissemination patterns over time among different Italian regions shows that the pre-lockdown phase was characterized by an exponential growth of the number of daily-confirmed COVID-19 cases and deaths, with highest incidence in the Northwest, followed by a significant decrease across all regions in the aftermath of lockdown measures (**Supplementary Figure 2)**. By the end of August 2020, epidemiological data also show increased and sustained transmission in the South and Insular regions, possibly driven by interregional spreading through small family/social network clusters. These regions are the main touristic destination for Italians, and most of the restrictions on international travel were still in place during Phase III ^16^. Lineages proportion and regional-specific distribution in different parts of the country are indicative of several independent founder events (**Figure 2B**). For example, lineage A, predominant in Sicily, has been detected in epidemiologically linked transmission chains that appear to be related to immigrants arrived from North Africa during the late Phase III ^17^. Interestingly, the number of circulating lineages have changed over time (**Supplementary Figure 3**). Sub lineage B.2 was the first one identified in January, marking the primary introduction of imported cases from China (no shown in Figure 2). Between February and April, additional sub lineages, such as B.1, B.1.1, and B.1.5 emerged in Northern and Central Italy, the epicenter of the first epidemic wave, likely reflecting subsequent importations. At the beginning of Phase II lockdown in May, which followed a dramatic decrease in cases, only B.1. and B.1.1 sub lineages were detected. During the period of epidemic recession between June and July, multiple sub lineages co-circulated again. However, the subsequent second wave was dominated by B.1.1 (September) and B.1. (October) (**Supplementary Figure 3**). Since Phase II and III measures permitted intra- and then inter-regional travel, respecitively, while country borders remained mostly closed (except with European countries part of the Shengen agreement), it is plausible that in the first epidemic wave lineages’ heterogeneity resulted from intial founder events associated with international travel, and then propagated through within state mobility during epidemic recession. Such sequence-based inferences, however, should be interpreted with caution because of the inherent sampling bias in SARS-CoV-2 full-length genomes currently available from Italian patients, which could affect results and limit their generalizability ^18^.

**Figure 2.**
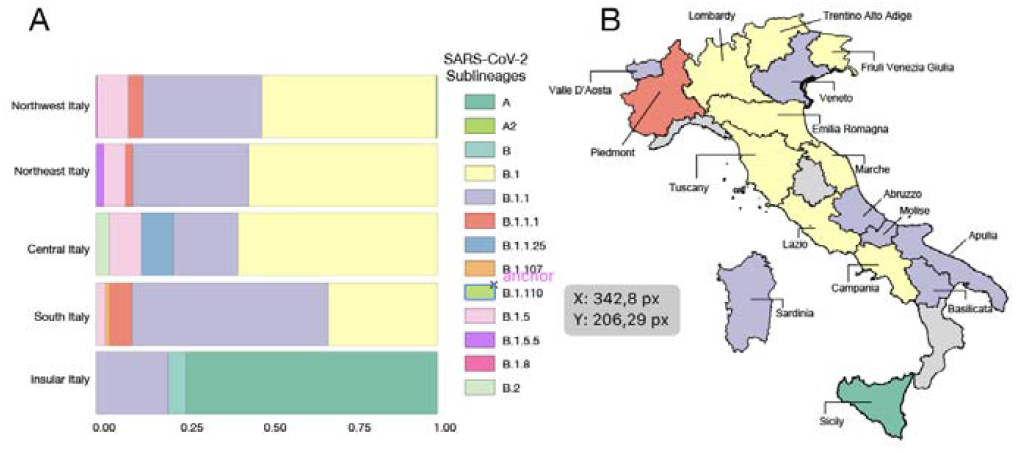
Frequency and distribution of SARS-CoV-2 lineages and sub lineages in Italy. **A**) Frequency of the lineages and sub lineages of SARS-CoV-2 among Italian macro regions. **B**) Distribution of the most prevalent lineage and sub lineage across the country.

**Figure 3.**
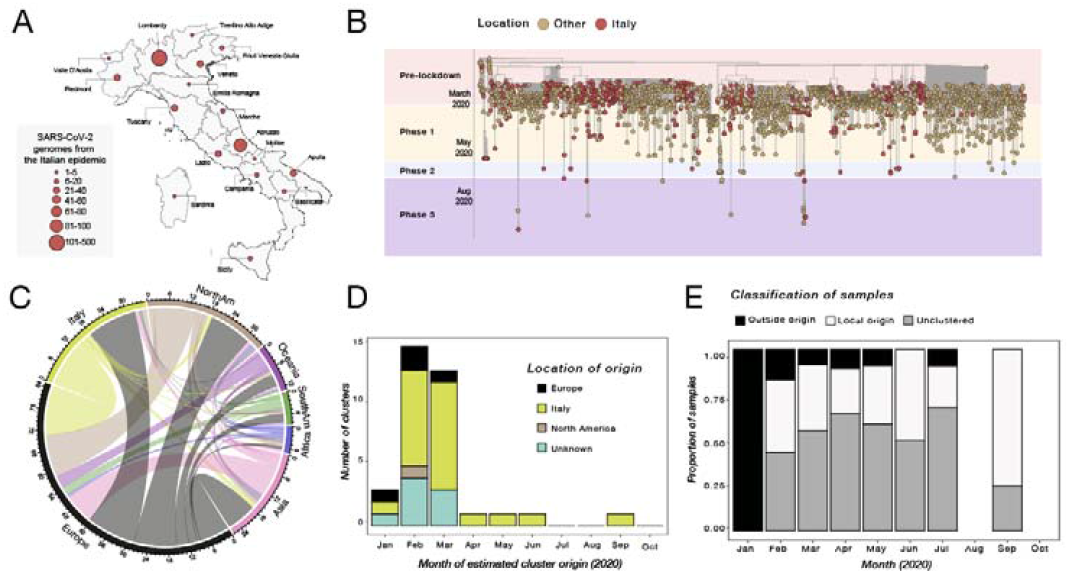
Phylogenetic characterization of Italian SARS-CoV-2 sequences. A) Map of Italy showing the number of SARS-CoV-2 genome sequences by region. The size of the circles indicates the number of new genomes available since the beginning of the epidemic in Italy; B) Time-resolved maximum likelihood tree of 1421 SARS-CoV-2 sequences including 714 from Italy (red circles). C) Chord diagram of estimated numbers of migration flows between the geographic areas. D) Frequency of estimated geographical origins for identified transmission clusters involving Italy and originating in the months of January through October of 2020. E) Frequency of Italian sequences (sampled from January through October) classified as unclustered (grey) or belonging to clusters with Italian (white) or non-Italian origins (black).

In our sequence dataset, only Lombardy (Northwest, most affected region so far), has provided a robust number of viral genomes (n=405), which in turn corresponds approximatively to one genome available every 450 positive cases. Abruzzo in Central Italy is the second most represented region in terms of available genomes (n=87), while many other regions, including Liguria (Northwest), Umbria (Central) and Calabria (South) are not comprehensively represented (**Figure 3A**), thus affecting our ability to characterize in-depth SARS-CoV-2 molecular epidemiology at a regional level. Nevertheless, phylogeny-inferred virus evolutionary patterns are useful to corroborate epidemiological data, test hypotheses regarding factors driving epidemic dynamics, and assess public health interventions such as stay-at-home orders. To this end, we time-scaled the best 100 ML trees of all available SARS-CoV-2 full genomes from Italian patients, and inferred the most likely location of each internal node (ancestral sequence) in the trees (see Methods for details). The overall topologies of the inferred trees were highly similar, and linear regression of root-to-tip genetic distances against sampling dates indicated sufficient temporal signal in the sequence data (**Supplementary Figure 4**). Although SARS-CoV-2 evolutionary rate in Italy was somewhat lower (1.44 10^−04^ nucleotide substitutions/site/year) than values obtained for the worldwide epidemic ^19,20^, the most recent common ancestor (TMRCA) of the available Italian sequences, ranged between January 2^nd^ and January 26^th^ (mean Jan 14^th^) 2020, consistent with the date of the first confirmed case (Jan 30^th^). Similarly, the root node (origin) of a time-scaled ML tree including both Italian (n=714) and worldwide reference sequences (n=1,421) was placed in China (99.8% probability), with a TMRCA dating back to early December 2019, in agreement with available epidemiology data ^21,22^, further validating our phylogeny inference. The tree (**Figure 3B**) consistently shows most of the Italian sequences interspersed with virus strains collected in other countries. This pattern, alike the one observed elsewhere ^23^, confirms that emergence of SARS-CoV-2 strains during the first epidemic wave was primarily fostered by travel exposure during the pre-lockdown phase, rather than interregional spreading. According to the estimation of migration flows, we further examined the potential Italian role as an exporter of SARS-CoV-2. The number of state transitions into and from Italy (**Figure 3 panel C**) heavily relies on the number and nature of the sequences that are included from other locations. Independently of the dataset, and in line with the epidemiological information, most of the geographical sources of the introductions are attributed to Europe (**Figure 3C**). Well supported (bootstrap values > 90%) putative transmission clusters within the phylogeny were identified based on a pre-defined genetic distance threshold likely to detect epidemiologically linked sequences (see Methods). Clusters containing at least one Italian sequence were considered of interest for the estimation of temporal and spatial origins of the transmission. Temporal origins of each cluster were derived from the clock-estimated age of the MRCA of all sequences belonging to the cluster. Spatial origins were inferred using joint likelihood ancestral state reconstruction, given known country of sampling of tip nodes (sampled sequences) within the tree. As expected, the number of (well supported) clusters formed over the course of the epidemic was largely influenced by the number of contemporaneous samples (**Figure 3D**), limiting conclusions regarding the rate of cluster formation over time. The estimated geographic origins of each cluster reflected the distribution of samples among the reference sequences, largely limited to Europe and North America. However, after April, transmission clusters could only be traced back to Italy, suggesting highly localized transmission following the implementation of Phase one lockdown measures. Each Italian sequence was then classified either as unclustered (i.e. no cluster with any other sequence with bootstrap >90%), or belonging to a local (all Italian) cluster. Italian sequences within well supported clusters including and originating from non-Italian strains were classified as belonging to “outside” clusters. Finally, each well supported cluster for which a single country could not be assigned with >90% probability as the one at the origin of that cluster, was also considered to be an outside (albeit unknown in origin) cluster. This revealed distinct patterns between January, February-July, and August (**Figure 3E**). All Italian sequences obtained in January belonged to clusters of foreign origin, demonstrating the influence of outside introductions before lockdowns were put into place. The predominant fraction was quickly replaced by sequences belonging to clusters of local origin and unclustered sequences, which suggests potential undersampling. The month of September, when the second epidemic wave was increasing, sequences of local origin, with no sequences of foreign origin, largely dominated. The fraction of sequences sampled in August (75%) was outside the 95% confidence interval (∼50%) for the fraction in remaining months, emphasizing the significant contribution of local transmission on sequences sampled in September. However, the specific mutational profile of the Italian sequences (**Figure 4A**), relative to the Wuhan reference (NC_045512), also provided some evidence of recently imported strains during Phase III lockdown, when travel bans began to be eased. In particular, 97.34% (n=695) of the available sequences carried the mutation encoding for the amino acid change D614G (genomic coordinate: *23403A>G*) in the Spike protein of SARS-CoV-2, while the remaining 2.66% (n=19) sequences displayed the nucleotide sequence encoding for the D614 wild type. The D614G mutation has been associated with higher infectivity and greater transmissibility with no effects on disease severity outcomes ^24–26^, although some of these findings have recently been questioned ^27^. The frequency of the D614G polymorphism among Italian regions over time (**Figure 4B**) shows that while G614 quickly became rapidly dominant during the first epidemic wave, and remained the only one detected in the available sequences through the first two lockdown phases. Thereafter, the D614 variants re-emerged following the relaxation of Phase III measures in Sicily (Insular Italy), possibly due to the epidemiologically-linked transmission chains related to immigration flow from North Africa ^17^, a scenario reinforced by SARS-CoV-2 lineage A prevalence in that region (**Figure 2B**).

**Figure 4.**
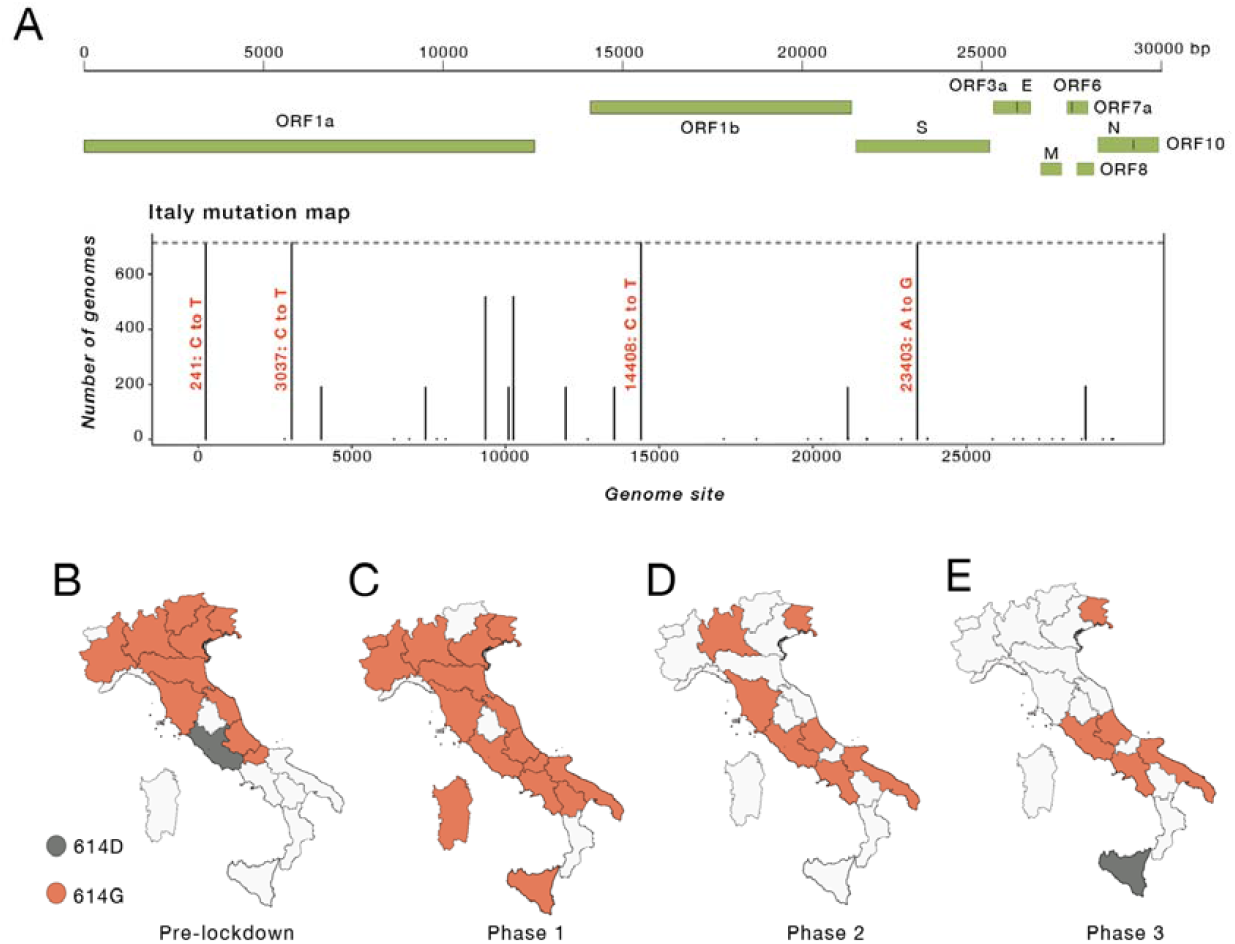
Italian strains mutations pattern. A) Variant maps of the most common mutations mapped against the SARS-CoV-2 genomes. Most common mutations defined as mutations present in >90% of the genomes in that group (black lines). B-E) Change in frequency of the D614G mutation in the Spike Protein across Italian regions during epidemic phases.

Results of cluster analysis indicate maintained local transmissions fostered by relatively small transmission chains during the months of low case reports and through the beginning of the second wave. This observation suggests that epidemic resurgence was associated with a relaxation of lockdown measures that led to increased local transmission, rather than a large number of virus re-introductions into the country. Such a scenario is also supported by surveys showing a significant reduction in the number of foreign tourists (about −65.9%), but an increase, albeit small (1.1%), of domestic tourism during the summer season after restrictions on interregional travel were relaxed ^16^. In order to explore whether increased mobility could explain the second surge of cases in Italy, we carried out stochastic agent-based epidemic simulations. Mobility data across three different modes of transportation (walking, public, and personal vehicle), derived from Apple Mobility trends reports, were used as a proxy for the number of individuals with whom an infected individual comes into contact, which was allowed to vary over time (see Methods). As the number of hospitalizations also dropped drastically (and stayed low) following the first surge in cases, the role of removal of infected individuals from the population *via* hospitalization was also tested, by allowing the probability of an infected individual exiting the simulation to be proportional to the standardized hospitalization rates (also varying in time). The simulated number of active infections over time using the mobility data alone, hospitalization data alone, and combined were then compared to the empirical case data. Whereas all three models produced a similar rate of new infections during the first epidemic wave (**Supplementary Figure 5**), time-varying rate of removal based on hospitalization rates (without mobility data) produced a continuing exponential growth of infections (**Figure 5A**). As in empirical data, time-varying number of contacts based on mobility produced two distinct waves, which were the most similar to the epidemic curve (**Figure 5B**). The model incorporating both mobility and hospitalization rates produced a first wave that was of too large a magnitude and a second wave too early in origin than the previous model (**Figure 5C**). The model incorporating mobility data alone resulted in the lowest mean absolute error (**Figure 5D**), producing a first wave of similar timing and magnitude and a delayed second wave, closer to the empirical epidemiology data (**Figure 1B**).

**Figure 5.**
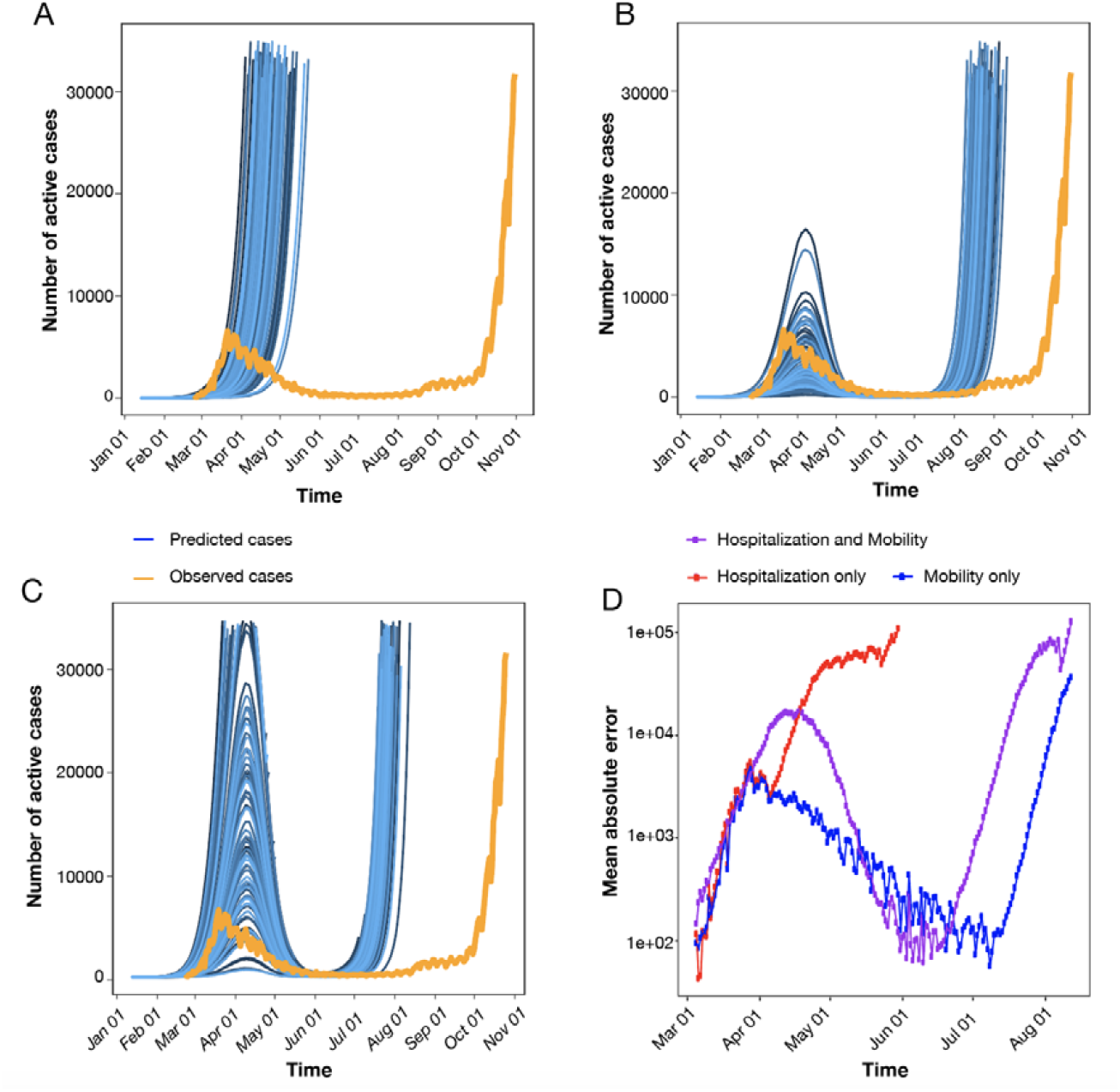
Simulated epidemics under scenarios involving time-varying mobility and hospitalization rates. A) The probability of removal of an infected individual was proportional to the empirical rate of hospitalization in the simulation of active infected cases over one year (blue). B) The number of individuals with whom an infected individual comes in contact was proportional to the empirically determined number of individuals utilizing walking, as well as public and personal modes of transportation, as primary means of mobility in the simulation of active infected cases over one year (blue). C) Hospitalization rates (as in A) and mobility data (as in B) were combined in the simulation of active infected cases over one year (blue). In A-C, orange represents the number of empirically observed infections. D) The absolute error was calculated for each time point of collected observations between simulated infections using hospitalization rates only (red), mobility data only (blue), and the combination (purple). Mean absolute error was calculated as the averaged error across 1,000 simulations. Simulation assumed a single outside introduction.

While results indicate that mobility data could reproduce epidemic wave patterns in Italy, we cannot exclude additional factors, that might have played a role delaying the second epidemic wave, not fully captured by our simulations, such as retention of restriction measures, different “types” of mobility between first and second wave, or higher temperatures in the summer ^28^.

By coupling phylodynamic analysis of viral genetic and epidemiology data, we show how the interplay between public health intervention and shifting SARS-CoV-2 transmission dynamics in Italy may explain the oscillation between times of relatively stable epidemic recession and dramatic resurgences, as it is currently being observed. This pattern of “rubberbanding” or “snapping back” after public health restrictions are lifted has, unfortunately, been followed by several other European countries. Overall, we show the critical role played by small transmission clusters, acting as “hidden reservoirs” during epidemic recession following aggressive lockdown measures, in maintaining SARS-CoV-2 low-level circulation in Italy, which eventually seeded a new epidemic wave. Despite the consistent agreement between different viral phylogeny-based and epidemiology data analyses, however, limitations of our work need to be acknowledged. Availability of a large number of viral sequences, collected over an extended period of time and sufficiently representative of the ongoing epidemic, is crucial for prompt genomic surveillance, and the evaluation and planning of effective and opportune control strategies. The number of Italian SARS-CoV-2 full genomes currently deposited in public databases represents a very small fraction (0.05%) of the documented number of confirmed cases in Italy, and sampling bias across regions differently affected by the epidemic further limits generalizability of the results. Moreover, our definition of putative transmission clusters (see Methods) does not require the sampling and inclusion of all the strains involved in a transmission chain, although it does allow for detection of monophyletic clades that likely comprise sequences epidemiologically linked through a transmission chain, whilebeit not directly. Nevertheless, epidemiology observations, corroborated by phylodynamic analyses based on available sequences, depicted a coherent picture. The first epidemic wave in Italy appears to have largely been linked to outside introductions leading to large transmission clusters, concomitant with high number of infections. Subsequent implementation of a three-phase nationwide lockdown strategy greatly mitigated numbers of infection and hospitalization during summer 2020. Yet, once mobility increased and social distancing decreased due to the progressive easing of lockdown measures, a sudden spike of infectious cases was observed, promptly followed by new hospitalizations. Our agent-based mathematical model recapitulates this phenomenon, further supporting the hypothesis that the small clusters observed during the summertime were acting, essentially, as “hidden reservoirs” that likely merged following the increased in mobility and reduction of social distancing measures. This in turn provided the “spark” for the sudden increase of infections observed at the end of summer, which led to the subsequent second wave of exponential grow. In other words, the drivers of SARS-CoV-2 transmission dynamics shifted from high levels of community transmission, likely involving mass super spreader events, in the early Italian epidemic, to sustainment by smaller family/social network clusters later in the epidemic. Unfortunately, this also suggests that no amount of community level interventions may be sufficient to curb the epidemic as long as people do not adhere to individual level measures such as mask use, hand hygiene, and social distancing. New lockdown measures are likely to provide only temporary relief, as has already happened in the first months of the epidemic in Italy and many other countries. Indeed, an important debate is currently ongoing about vaccine deployment, given financial and logistic restrictions mandating a very long phased deployment, based on prioritization policies. In this context, our results suggest that hidden transmission reservoirs may continue to sustain local outbreaks into late 2021, as vaccine rollout will likely take months before reaching the necessary herd-immunity threshold. Ultimately, our ability to curb successfully the current pandemic, may be linked to our ability to determine number and structure of such reservoirs within the social and behavioral context of specific locales.

## Data Availability

Sequence and epidemiology raw data utilized, generated or analyzed during these studies are available from the authors upon request (including sequence alignment and R scripts for the phylodynamic analyses).

## Acknowledgments

We thank all the authors, originating and submitting laboratories that have kindly deposited and shared genome data on GISAID EpiCoV database, on which this research is based. An acknowledgment table can be found in **Supplementary Table 3**. We thank all personnel from the Italian Health Surveillance System of the *Protezione Civile* that assisted with epidemiological data collection. MS, BDR and CB contribution was funded, in part, by the U.S. National Science Foundation RAPID grant (DMS-2028728). MS and BDR are also supported by the Stephany W. Holloway University Chair in AIDS Research.

## Authors’ contributions

Conception and design: MG, EC, BRM, MS and MC; Data collection: MG, EC; Investigations: MG, EC, FB, BRM, VF, SF, and JL; Data Analysis: MG, EC, BRM, EW, VF, NED, CB and JL; Writing – Original: MG, EC, FB, BRM, MS, DZ and MC; Draft Preparation: MG, EC, FB, BRM, JL, VT, MS, DZ and MC; Revision: GC, SP, AR, AB, VT, CS, AM, TA, EW, TdO, LCJA, GC, RC, AC, JL, MS, DZ, and MC.

## Methods

### Sequence data collection

To perform a comprehensive analysis of the genomic epidemiology of SARS-CoV-2 in Italy, after excluding low-quality genomes (> 10% of ambiguous positions), we dowloaded all Italian full-length viral genomes available on GISAID (https://www.gisaid.org/) (n=714) up to October 31^h^ 2020. Appropriate acknowledgement was given to the sequencing laboratories (**Supplementary Data S1**). Sampling locations of available genomes in this dataset included 17 of 20 regions in Italy, and collection dates spanned from January 30^th^ (the first two imported cases in Italy) to October 1^th^ 2020. Each Italian sequence was used in a local alignment (BLAST) ^29^ search for the most (genetically) similar non-Italian sequence in the GISAID database as of Oct 31^st^, 2020, and linked to two reference sequences including the best match (highest E-value) with a date occurring within one month following, as well as one month prior to the sampling date of the Italian sequence (although, in some cases, only a single non-Italian reference sequence fulfilling one of the inclusion criteria could be found for multiple Italian query sequences). After removing duplicate sequences and masking mutations potentially associated with common sequencing errors, using a vcf filter ^30^, a final dataset of 1,421 reference sequences was assembled (**Supplementary Table 2**).

### Sequence alignments and phylogenetic analysis

Sequences (Italian + reference strains) were aligned using MAFFT (FF-NS-2 algorithm) employing default parameters ^31^. The alignment was manually curated to optimize number and location of gaps using Aliview ^32^. A site-specific mutational comparison of the 714 Italian genomic sequences obtained from the GISAID database was made with the MAFFT-aligned SARS-CoV-2 reference genome (RefSeq: NC_045512.2), obtained from the GenBank database. Lineage assessment was conducted using the Phylogenetic Assignment of Named Global Outbreak LINeages tool available at https://github.com/hCoV-2019/pangolin ^33^. Phylogenetic analysis of was performed using the maximum likelihood (ML) method implemented in IQ-TREE (version 1.6.10), employing the best-fit model of nucleotide substitution according to the Bayesian Information Criterion (BIC), as indicated by the Model Finder application implemented in IQ-TREE ^34^. The statistical robustness of individual nodes was determined using 1000 bootstrap replicates.

### Molecular clock calibration and estimation of virus effective population size

ML trees were inspected in TempEst v1.5.3 for the presence of temporal signal (i.e., linear relationship between genetic distance and sampling time in the available sequences) [21]. The treedater package in R v3.6.0 ^35,36^ was used for molecular clock calibration of the Italy-only data, as well as the combined Italy and reference data. The top 100 maximum likelihood (ML) trees (i.e. the trees with the 100 lowest -*log*[likelihood] values), were chosen for calibration according to a strict clock (no branch specificity) among the Italy-only data, whereas a single-best ML tree was chosen for the combined dataset. Individual *taxa* sampling times were used to rescale branch lengths to time in each tree using a starting value of 8×10^−4^ substitutions/site/year. The skygrowth non-parametric demographic model ^37^ was then used in R with time-scaled trees to estimate of median virus effective population size (*Ne*) and 95% high posterior density intervals for each week during the epidemic in Italy (Italy-only dataset) using the default smoothing parameter value (tau) of 0.1.

### SARS-CoV-2 transmission cluster identification and characterization

Transmission clusters were identified using Phylopart v2 ^38^ applied to the ML tree of combined sequence data (scaled in substitutions/site). A range of percentile thresholds spanning 10^−6^ % – 15% of the whole-tree patristic distance distribution was used to choose an optimal threshold point and to verify robustness of cluster composition. The minimum percentile threshold that maximized the number of clusters was chosen as the optimal threshold by performing multiple clustering runs on randomly sampled patristic distance distributions (1 million for each run). Well-supported sub-trees (bootstrap values > 90%) with mean pairwise patristic distances among *taxa* within the chosen threshold were considered putative transmission clusters (i.e. clusters comprising sequences epidemiologically linked through a transmission chain, although some of the direct links may be missing). Only clusters containing at least 1 Italian sequence were considered in downstream analyses. The phytools package ^39^ in R was used for joint likelihood reconstruction of discrete ancestral origins ^40^ according to country (and associated uncertainty) for the most recent common ancestor (MRCA) of each transmission cluster within the ML tree (scaled in substitutions/site) for the combined dataset. Transition rates among discrete states (countries) along tree nodes were considered to be equal. The tree scaled in time was used to attribute temporal origins to each cluster, or time of MRCA (TMRCA). The following R packages were used in the manipulation of data for cluster characterization and visualization: ape ^41^, dplyr ^42^, purr ^43^, rlist ^44^, tidytree ^45^, ggplot2 ^46^, data.table ^47^, reshape2 ^48^, lubridate ^49^, ggtree ^50^, tidyr ^51^, parallel ^36^.

### Estimation of basic reproduction number

Estimates for daily basic reproduction number, *Re*, of SARS-CoV-2 in Italy were obtained from the COVID-19-re data repository (https://github.com/covid-19-Re/dailyRe-Data) as at 20^th^ September 2020. The effective reproductive number describes the average number of secondary infections caused by an infected individual. The relevant method of calculation of Re builds upon another method developed by Cori et al.^52^, accessible through EpiEstim R package. Instead of using a time series of infection incidence, which cannot be observed directly, the relevant method infers the infection incidence time series based on secondary sources of information such as COVID-19 confirmed case data, hospital admissions, and deaths. This was considered in combination with two other sets of time variables: i) the duration of SARS-CoV-2 incubation period and ii) the time delays between the onset of the symptoms and a positive test, a hospital admission or the death of a patient. The relevant method infers infection time series from the stated observed incidence data by deconvolution ^52–55^.

### Epidemiology data assembly

We analysed COVID-19 cases counts in Italy from publicly released data up to October 31^st^ 2020 from the Italian Civil Protection Department repository (https://github.com/pcm-dpc/COVID-19) that releases daily updates on the number of new confirmed cases, deaths and recoveries, with a breakdown by region. To illustrate the epidemic progression, the daily number of confirmed cases of people infected with SARS-Cov-2 in Italy was plotted alongside a timeline of lockdown phases and variation in estimated virus reproduction number until October 31^st^ 2020. For convenience the geographical locations were aggregated by Italian macro regions: Northeast, Northwest, Central, South, and Insular, which are basic regions for the application of regional policies (Italian regions). Mobility data over time, combining data on three different forms of transportation - personal vehicle, public, and walking - were obtained from Apple Mobility Trends Reports (https://covid19.apple.com/mobility).

### Agent-based stochastic model simulation of the Italian epidemic

The Italian epidemic was simulated using the forward-time, agent-based stochastic transmission chain simulator, nosoi ^56^, which allows for time-varying parameterization. The simulation was initiated with a single infected individual with a probability of transmission of 0.02 per day following an incubation period, which was set to a mean of 5 days and standard deviation of 2 days. The rate of transmission was fixed throughout the simulation at this value. Following the incubation period, each individual was considered infectious for approximately 9 days, exiting the simulation at a mean time of 14 days (standard deviation equal to 2) after infection. Three different simulation scenarios were tested, assuming a direct relationship between 1) rate of mobility and the number of other individuals with which each infected individual comes into contact, 2) hospitalization rate and the rate at which an infected individual was removed from the simulation during the infectious period (approximately 5 to 14 days), or both. Hospitalization and mobility data were standardized by (observed-minimum)/(maximum-minimum) to a range of 0-1 and modeled using a Fourier series periodic function, with mobility data comprised of 2 sine/cosine terms (linear model regression R2=0.8484) and hospitalization data of 4 sine/cosine terms (linear model regression R^2^=0.984). In scenarios 1 and 3, the probability of exiting the simulation was allowed to vary over time proportionally to the standardized rate of hospitalization, resulting in a maximum of ∼35% of infected individuals hospitalized during peak hospitalization of the epidemic. In scenarios 2 and 3, the number of contacts for each infected individual was allowed to vary over time proportionally to the mobility rate, resulting in a mean of approximately 15 individuals in contact with each infected individual. For scenario 1, a static mean removal rate (during the infectious period described above) over time was set to 0.04 (standard deviation of 0.01). For scenario 2, the mean number of contacts per individual was set to 15 (standard deviation of 8). The mean absolute error for each time point was calculated to assess the deviation of the simulated number of actively infected individuals for each of the three scenarios from the true number of cases, provided by Italian Ministry of Health and the Civil Protection Department.

---

**Supplementary Figure 1.**
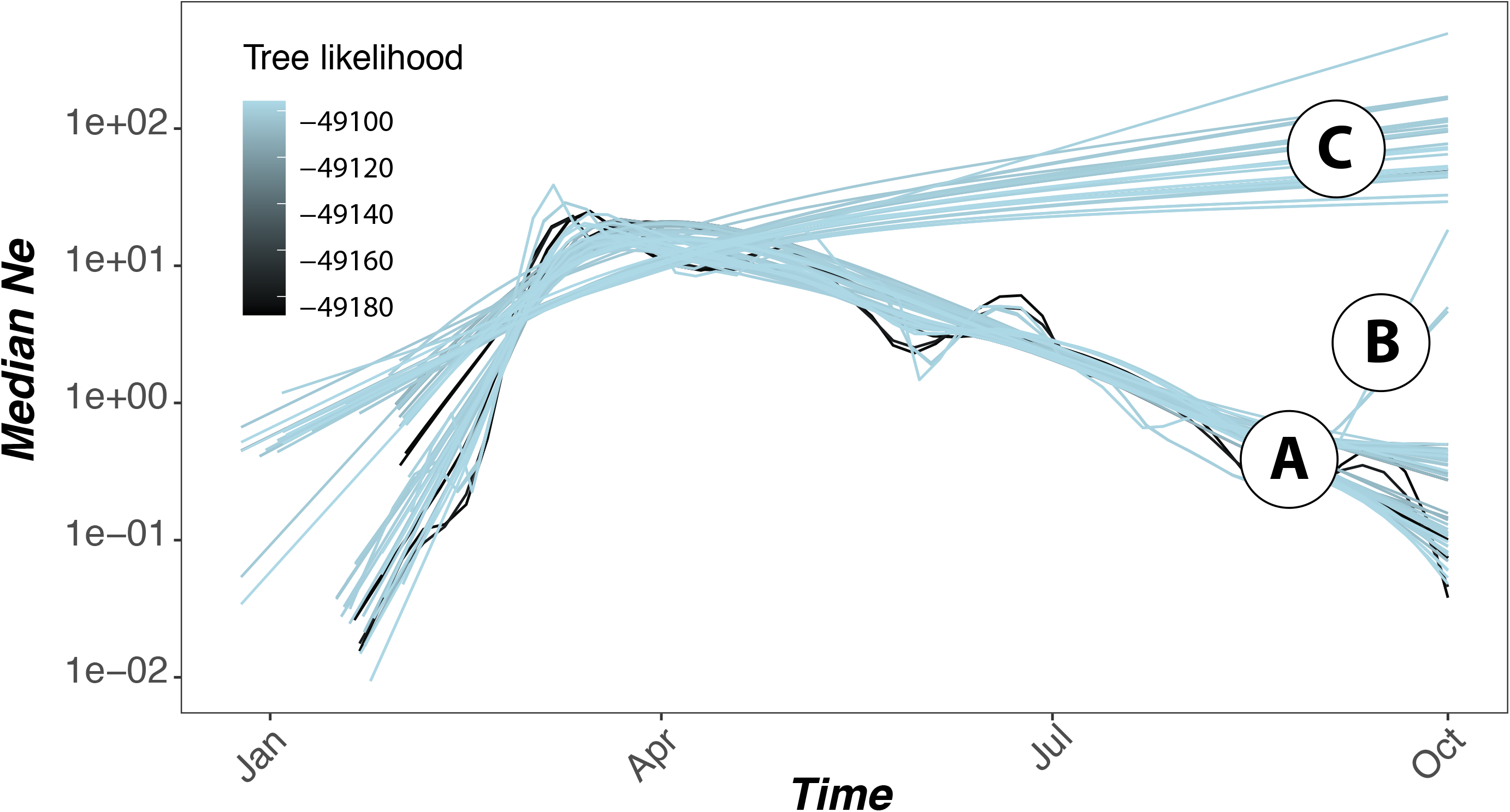
Estimates of the viral effective population size (Ne) in the Italian epidemic. Estimates of the viral effective population size (Ne) using a sample of 100 phylogenetic trees with the highest log-likelihood values. Encircled letters (A, B and C) label the three major patterns inferred from collection of trees with the highest likelihood values.

**Supplementary Figure 2.**
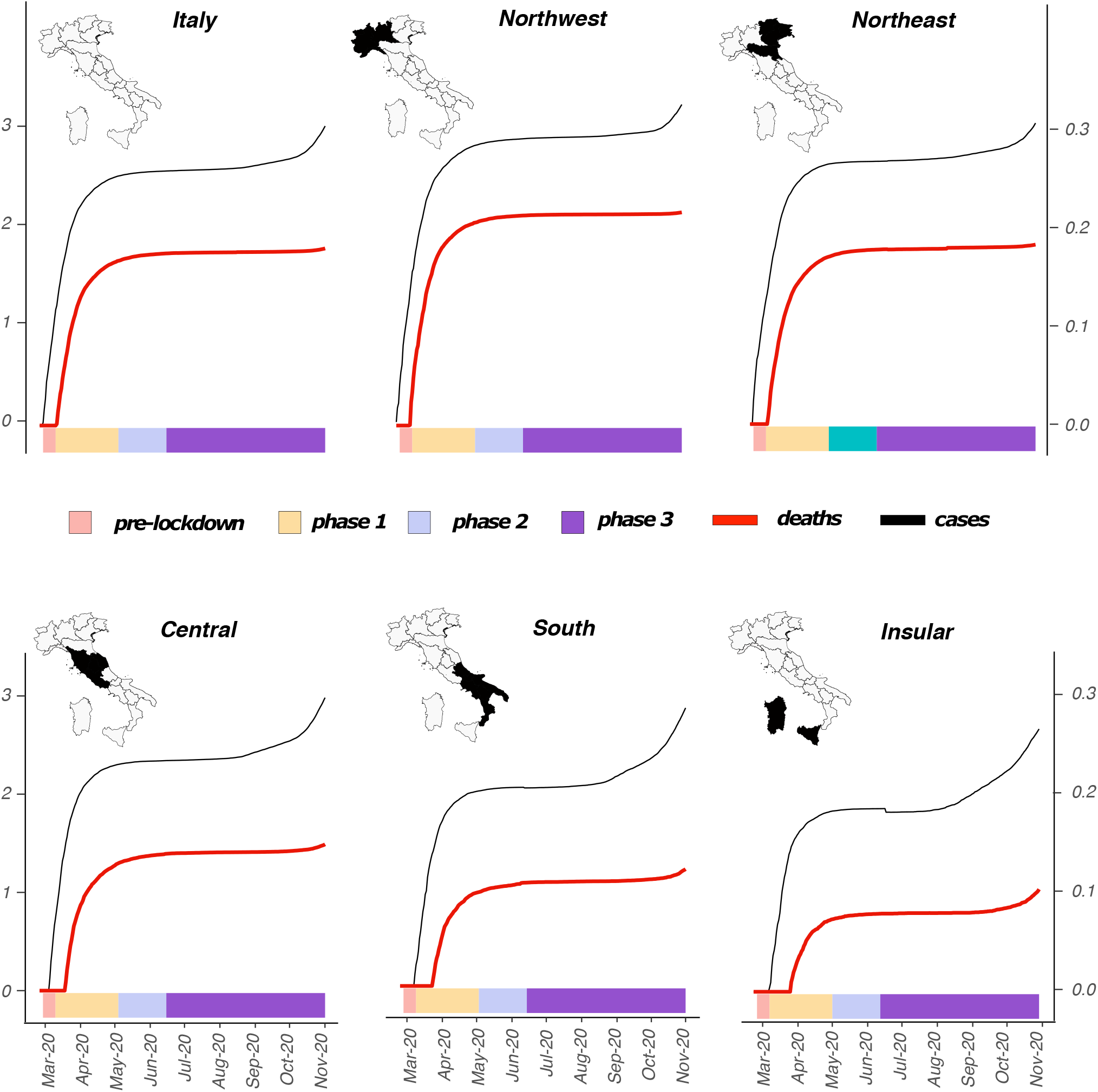
Spatial and temporal distribution of SARS-CoV-2 trough Italian regions. Italian regions were aggregated into five macro regions (NUTS, Nomenclature of territorial units for statistics): Northeast, Northwest, Central, South and Insular. The left-hand Y-axis (left) represents incidence (cases per 100K population, black curve) of COVID-19, while the secondary Y-axis (right) represents the number of deaths (red curve) related to COVID-19 through the country. Y-axis numbers are represented as log10 for visual purposes. Colours at the bottom represent the epidemic phases in Italy: pre-lockdown (early transmission) in light red, phase 1 in yellow, phase 2 in light violet and phase 3 in purple. Maps of Italy were superimposed to showing to exact location of each region: Northwest; Northeast; Central; South and Insular.

**Supplementary Figure 3.**
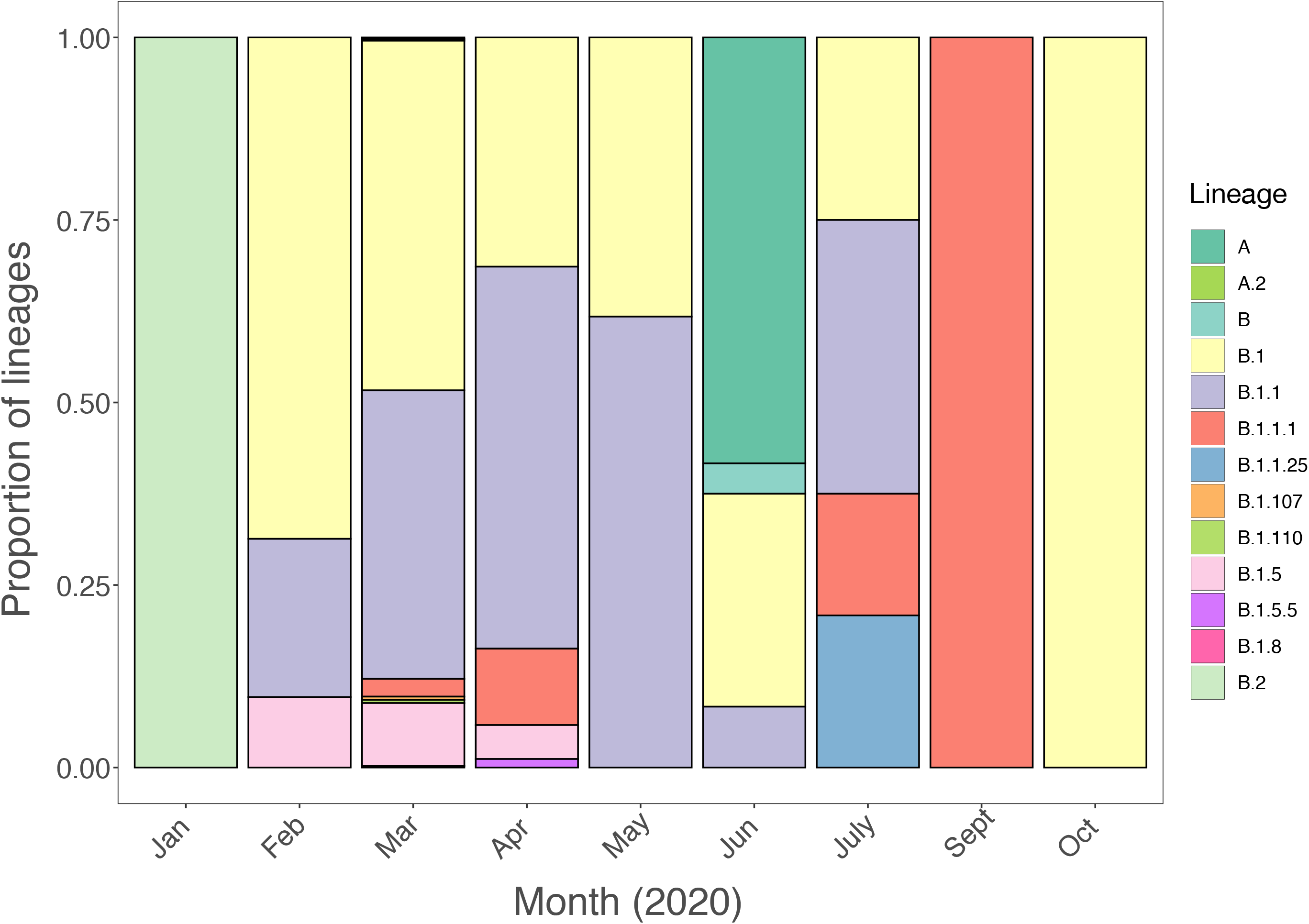
Frequency and distribution of SARS-CoV-2 lineages in Italy over time.

**Supplementary Figure 4.**
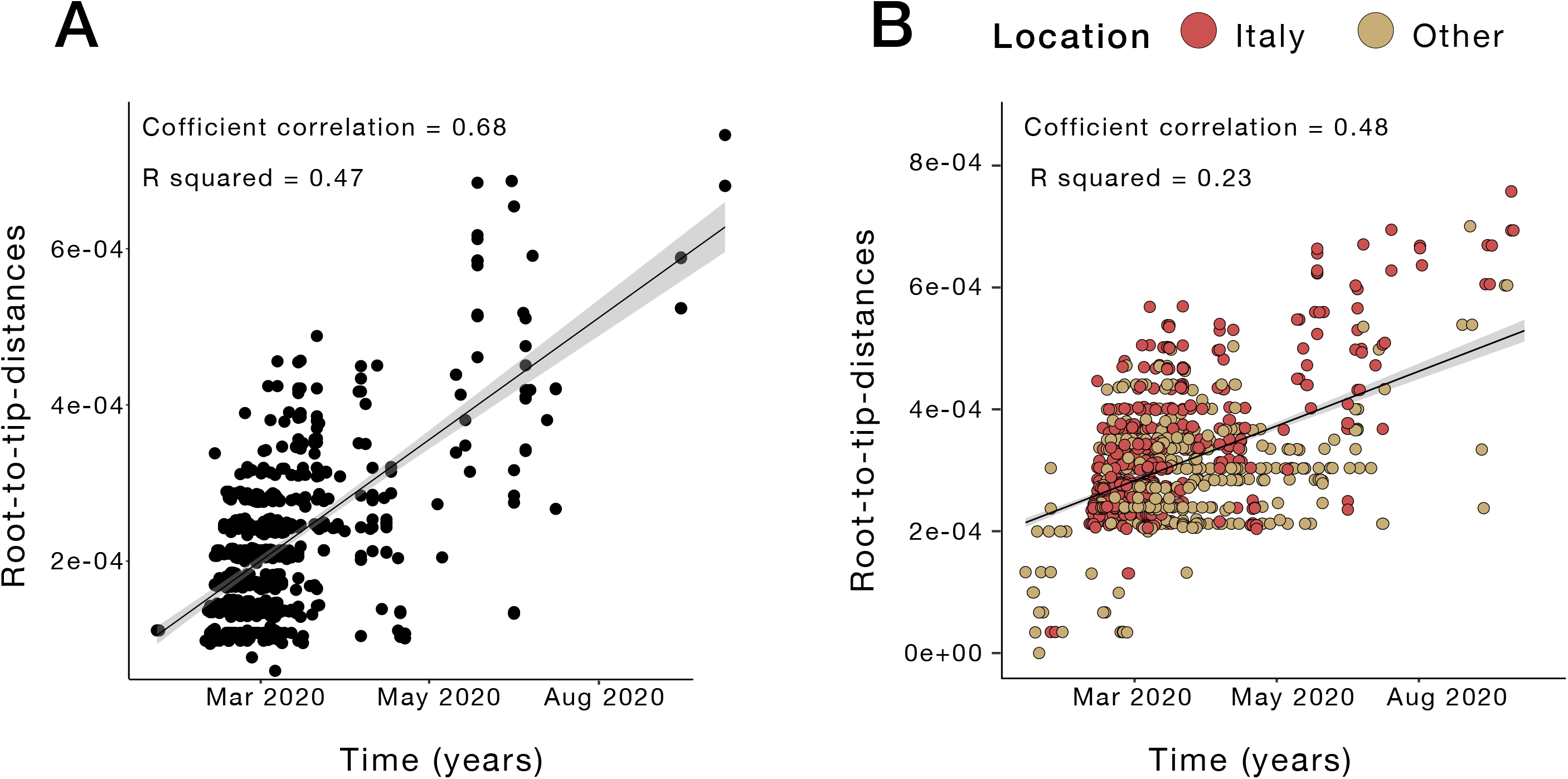
Analysis of temporal structure. **A.** Root-to-tip genetic divergence of Italian sequences against time of sampling. **B.** Root-to-tip genetic divergence for the whole dataset (Italian strains + reference sequences) against time of sampling.

**Supplementary Figure 5.**
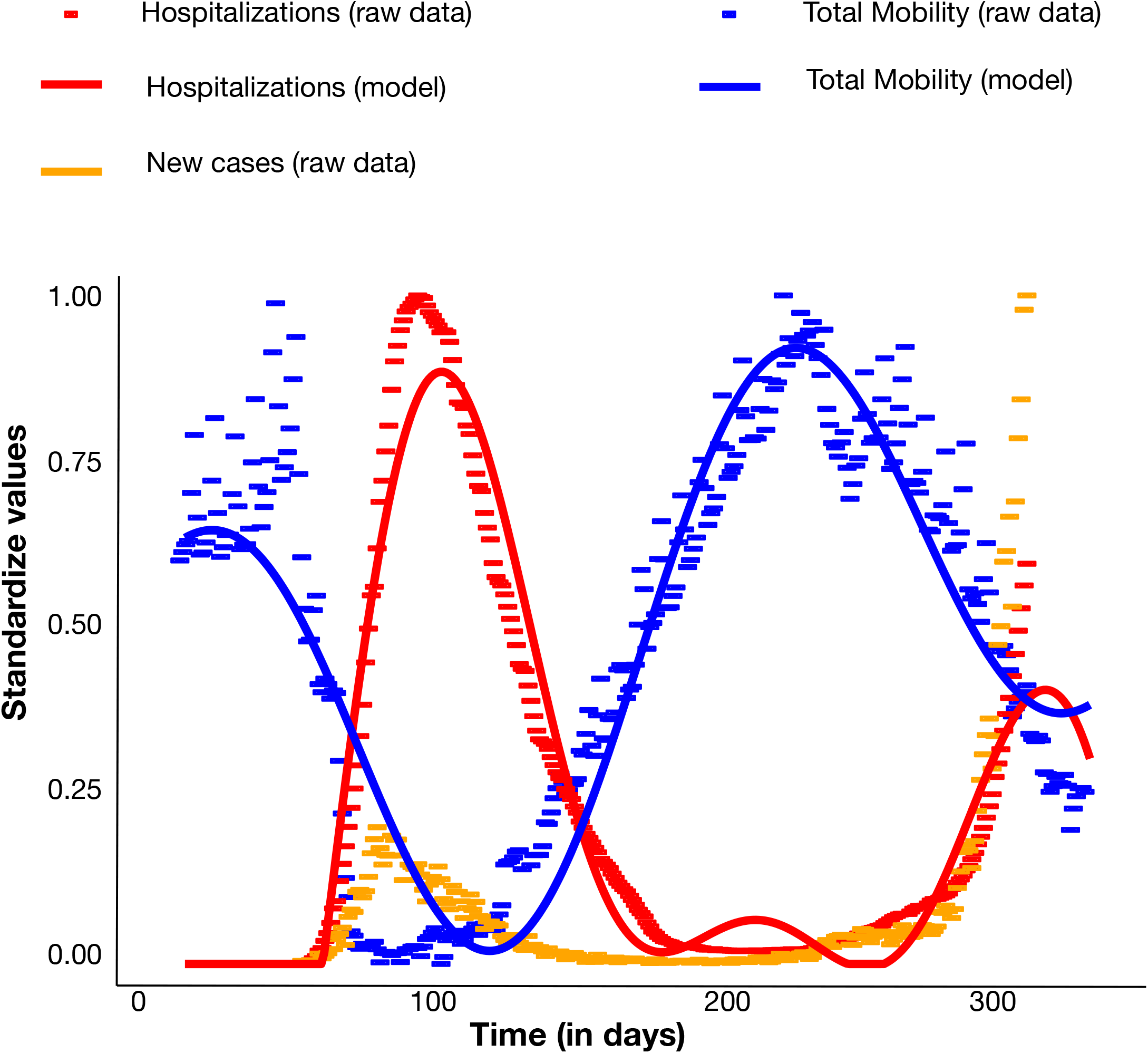
Empirical observations (points) and chosen model (lines) for hospitalization rates (red) and number of mobile individuals (blue) over time. Hospitalization rates represent the number hospitalized per positive case, whereas mobility represent the number of individuals utilizing walking, as well as public and personal modes of transportation, as primary means of mobility (blue). Values were standardized for comparison. Empirically observed number of new infections (standardized) are also shown for comparison in orange.

